# Two phenotypes of Chronic Recurrent Multifocal Osteomyelitis with different patterns of bone involvement

**DOI:** 10.1101/2022.05.10.22274859

**Authors:** Dita Cebecauerová, Hana Malcová, Veronika Koukolská, Zuzana Kvíčalová, Ondřej Souček, Lukáš Wagenknecht, Jiří Bronský, Zdeněk Šumník, Martin Kynčl, Marek Cebecauer, Rudolf Horváth

**Affiliations:** Department of Paediatric and Adult Rheumatology, Motol University Hospital, Prague, Czech Republic; Department of Radiology, Second Faculty of Medicine, Charles University and Motol University Hospital, Prague, Czech Republic; Department of Biophysical Chemistry, J. Heyrovsky Institute of Physical Chemistry of the Czech Academy of Sciences, Prague, Czech Republic; Department of Pediatrics, Second Faculty of Medicine, Charles University and Motol University Hospital, Prague, Czech Republic; Department of Ortopaedics, Second Faculty of Medicine, Motol University Hospital, Prague, Czech Republic

**Keywords:** Autoinflammatory disease, chronic non-bacterial osteomyelitis, chronic recurrent multifocal osteomyelitis, inflammatory bowel disease, sacroiliitis, whole-body magnetic resonance imaging

## Abstract

**Introduction:** Chronic Recurrent Multifocal Osteomyelitis (CRMO) is an autoinflammatory bone disorder with predominantly paediatric onset. Children present with multifocal osteolytic lesions accompanied by bone pain and soft tissue swelling. Patients often exhibit extraosseous co-morbidities such as psoriasis, inflammatory bowel disease, and arthritis.

**Objectives:** Comparison of children with two different phenotypes of CRMO defined by presence or absence of extraosseous co-morbidities.

**Methods:** Children diagnosed with CRMO at the Motol University Hospital between 2010-2020 were retrospectively reviewed, and according to the absence or presence of extraosseous manifestations divided into two cohorts – osteolytic CRMO and complex CRMO. The two groups were compared in terms of demographic data, age at disease onset, number and site of bone lesions, laboratory biomarker values, and need of escalation to a second-line therapy

**Results:** Thirty-seven children (30 female, 7 male) with confirmed CRMO were included in the analysis. The mean age at disease onset was ten years. All but 3 patients presented with multifocal disease. Twenty-three children (62%) had at least one extraosseous manifestation (13 sacroiliitis, 8 inflammatory bowel disease, 6 skin disease [acne, pustulosis, or psoriasis], 7 arthritis). Complex CRMO was associated with a significantly higher ESR rate (p=0.0064) and CRP level (p=0.018). The groups did not differ in number of foci or in age at disease onset. Bone lesion distribution differed between the two groups with significantly more frequent involvement of clavicle (p=0.011) and pelvis (p=0.038) in patients with complex CRMO. Children with complex CRMO more often needed escalation of therapy to DMARDs and biologic agents.

**Conclusion:** Our data suggest that CRMO affecting solely the skeleton has milder course compared to complex CRMO with extraskeletal features. Further studies are needed to explore the clinical as well as the patient reported outcomes and promote individually tailored therapeutic strategies in both CRMO phenotypes.

## Introduction

Chronic Recurrent Multifocal Osteomyelitis (CRMO) is a rare autoinflammatory disorder characterized by sterile bone lesions ^1,2^. It is often referred to as a subgroup within Chronic Non-Bacterial Osteomyelitis (CNO). For patients with concomitant acne or pustulosis the acronym SAPHO (Synovitis, acne, pustulosis, hyperostosis, osteitis) is used. CRMO incidence is estimated at 1:250 000 ^3^ ; precise values remain unknown. However, the number of diagnosed cases has increased lately with improved public and care-provider awareness and better accessibility of whole-body magnetic resonance imaging (WB-MRI) ^4,5^. It affects predominantly young girls with peak presentation at age 10 years ^6-8^, typically with bone pain accompanied by local warmth and soft tissue swelling. The disease course is recurrent or persistent with a variable number of osteolytic lesions developing over time. The unifocal and persistent form is more common in adult patients ^9^. Adults and adolescents often display involvement of clavicle and sternum and have various cutaneous manifestations ^9,10^.

The molecular pathogenesis is not fully understood, but there is a consensus that CRMO is an autoinflammatory disease, characterized by unprovoked spontaneous flares of tissue-specific bone inflammation. The fact that different autoimmune conditions frequently affect pedigrees of CRMO patients emphasizes the role of genetic predispositions. However, specific genetic variants are characterised only in a small proportion of patients ^11-14^. Typical examples of monogenic forms of osteomyelitis represent Majeed syndrome or DIRA (Deficiency of Interleukin-1 Receptor Antagonist), with an altered IL-1 signalling pathway. Mutations in the coding and regulatory parts of *FLBIM1* are found in a minority of children with non-syndromic CRMO together with an altered IL-10 signalling pathway ^11,12^. These variants are, nevertheless, considered as contributing to rather than as underlying CRMO. At this writing, no other entries for CRMO exist in the Online Mendelian Inheritance in Man database^15^. Clinically, CRMO shares significant overlap with TNFα-related autoimmune conditions such as spondyloarthritis, psoriasis, and inflammatory bowel disease (IBD). Indeed, CRMO and/or SAPHO syndrome may, over time, evolve into spondyloarthropathy in some patients ^16^. The available data are limited but suggest that aetiopathogenesis of CRMO is complex and that a probably heterogenous genetic background underlies a seemingly uniform phenotype.

Recent reports indicate that about half of CRMO patients have associated extraosseous involvement, mostly psoriasis and palmo-plantar pustulosis, IBD, and arthritis. The frequency of particular co-morbidities varies significantly among reported cohorts^6,8,9,17,18^. Whilst cutaneous involvement is reported in 10-30% of patients, IBD is diagnosed in a much smaller group (5-13%). Arthritis varies significantly between individual cohorts (6-56%). Evidently, more studies are required better to understand these complex relations between CRMO and associated conditions.

So far, no specific guidelines exist for the treatment of children with CRMO. Therapy response assessment is complicated by the lack of a definition of disease remission. Based on current treatment practices, Zhao et al. published consensus treatment plans for CRMO refractory to non-steroidal anti-inflammatory drugs (NSAIDs) ^10^. Whereas NSAIDs represent first-line treatment, bisphosphonates are used in case of spinal involvement or NSAIDs failure. Disease-modifying anti-rheumatic drugs (DMARDs) and short-course corticosteroids can be used to escalate therapy; however, disease remission is achieved in fewer patients for whom these approaches are required ^6,9,19,20^. Biologic DMARDs, primarily tumour necrosis factor α inhibitors (TNFi), are the preferred choice in refractory disease or associated conditions, including sacroiliitis or IBD ^21,22^.

This study postulated that the two distinct phenotypic manifestations of CRMO – a solely osteolytic form, with only bone involvement, and a complex form, characterised by the presence of extraosseous associated conditions – might differ in clinical and laboratory aspects of disease, and sought to compare cohorts of children with the two forms of CRMO.

## Methods

We retrospectively reviewed and analysed a cohort of paediatric CRMO patients diagnosed and treated at Motol University Hospital, Prague, between 2010 and 2020. Only children still followed-up in Motol University Hospital, were included in the study. The diagnosis was based on the typical clinical and radiological findings and in 62% of children was completed by histologic and microbiologic confirmation of chronic non-bacterial osteomyelitis. Bristol criteria^17,23^ were applied retrospectively to all patients included in the study. The study was approved by the local Ethics Committee (EK 1412/20). Written informed consent was obtained from patients or legal guardians before study enrolment. From medical records we collected clinical data, including family history, gender, ethnicity, age of disease onset, first symptoms at manifestation, number and distribution of bone lesions, biopsy results, microbiologic results, spinal involvement, inflammatory and immunological marker values, faecal calprotectin levels, extraosseous disease, and therapy. WB-MRI and X-ray imaging were re-assessed by an experienced radiologist. Sacroiliac-joint involvement was diagnosed only when typical oedema of subchondral bone was present.

### Statistical analysis

Fisher’s exact test was used to compare the two groups of patients with lesions present and absent at a given site. All datasets (age at onset, number of foci, erythrocyte sedimentation rate [ESR], and serum C-reactive protein [CRP]) were first tested for normality using quantile-quantile plots and the Anderson-Darling test. Number of lesions and age at onset showed normal distributions with equal variance (two-sample F-test). Two-sample unpaired Student’s t-testing was used to compare the means between the osteolytic and complex CRMO groups. For ESR and CRP parameters not following a normal distribution, means were compared with the two-sample Kolmogorov-Smirnoff test. Statistical analyses were performed using appropriate functions of MATLAB software (Mathworks, Natick, MA).

## Results

Thirty-seven children, 30 girls (81%) and 7 boys (19%), were enrolled in the study (Fig. 1). Their clinical characteristics are summarized in Tab. 1. The initial symptom in most cases was limb pain (51%), followed by back pain (38%). The prevalently affected site was pelvis (59%), followed by femur (49%), tibia (43%), spine (38%), and clavicle (35%). Clavicular swelling as a first sign was present in 2 patients (5%); however, WB-MRI revealed asymptomatic clavicular lesions in 11 more children, with bilateral involvement in 4. Diaphyseal regions were affected in 9 children (24%). In six the lesion laid within the large metaphyseal-diaphyseal region. Solely diaphyseal lesions were confirmed in 4 children (11%). The least involved regions were skull, mandible, and scapula, with involvement in 2 children each. Just 3 patients (8%) were proven to have a single osteolytic site on WB-MRI. These solitary lesions were in 10^th^ thoracic vertebra, clavicle, and humerus.

**Figure 1.**
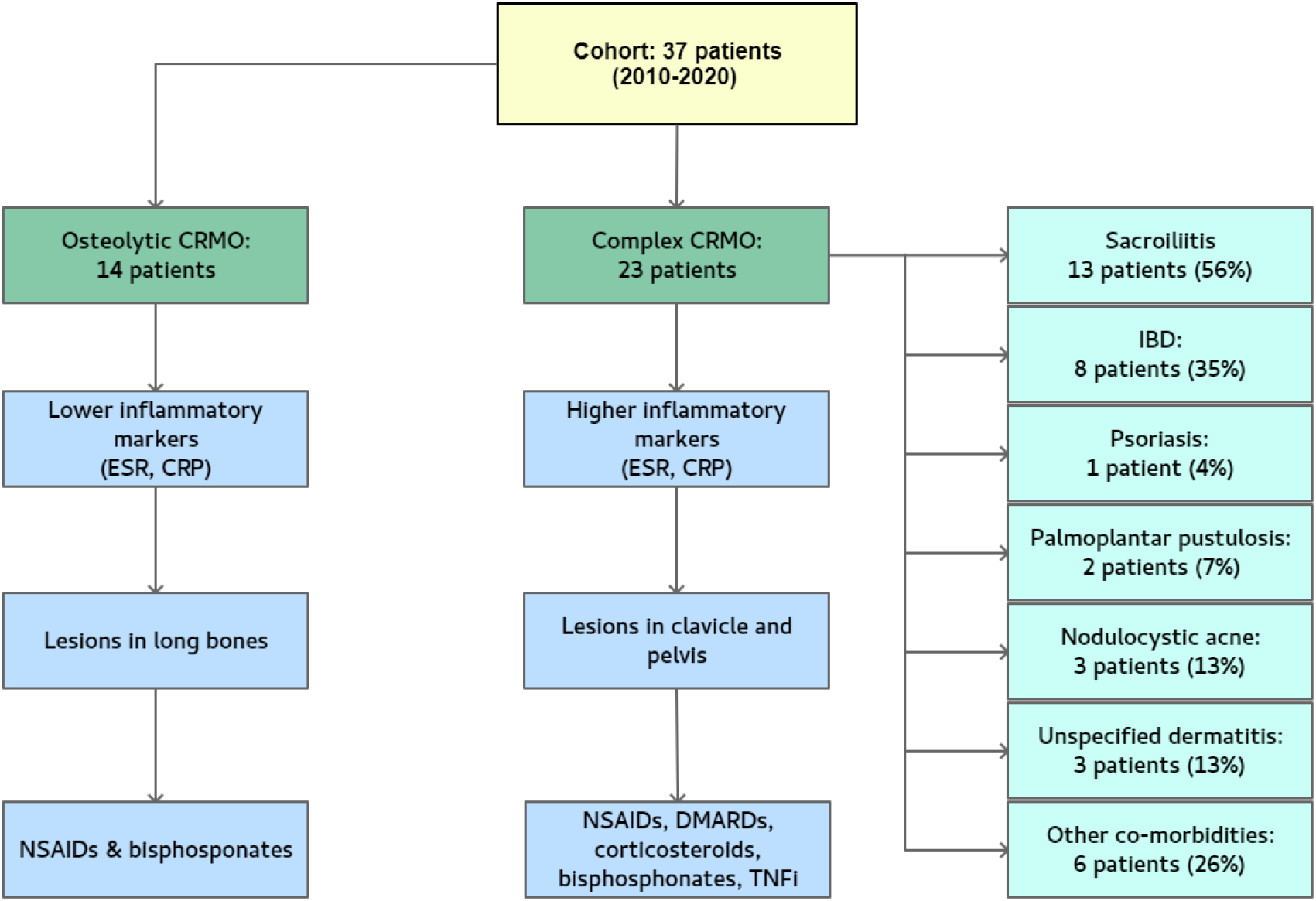
A graphical scheme summarising critical findings in our cohort. CRP – C-reactive protein, DMARD - disease-modifying antirheumatic drugs, ESR - erythrocyte sedimentation rate, IBD – inflammatory bowel disease, NSAID - non-steroidal anti-inflammatory drugs, TNFi – tumour necrosis factor α inhibitors.

Inflammation-marker values were elevated at presentation in most of the patients. ESR and/or CRP were increased in 24 children (65%) at the time of diagnosis, but they did not intercorrelate in all patients, *i*.*e*., some children had elevated ESR but not CRP and *vice versa*. Only 5 children had inflammatory-marker values within normal range. ANA were identified in 2 patients (5%). HLA-B27 positivity was detected in 6 children (16%), an incidence twice as high as that in a general Central European population ^24^.

Twenty-five children (68%) had family histories of one or more autoimmune diseases in first-and second-degree relatives (Fig. 2a), most often IBD (6) and psoriasis (3). Twenty-three CRMO patients (62%) had associated extraosseous conditions (Fig. 2b) at the time of diagnosis or developed them during follow-up (1-10 years). These were considered as ‘complex CRMO’ in this study. Thirteen children (35%) had sacroiliitis (Tab. 1), 7 of them symptomatic. In 6 patients, sacroiliitis remained clinically silent. Bilateral sacroiliitis was more frequent than unilateral and was not associated with HLA-B27 positivity. Only one HLA-B27 positive child had MRI-proved sacroiliitis (Fig. 3). In this patient, sacroiliitis remained asymptomatic despite ongoing inflammation detectable by MRI. On the other hand, 5 out of 6 HLA-B27 positive children had vertebral lesions (Fig. 3). Four children who were treated with bisphosphonates achieved regression of sacroiliitis.

**Figure 2.**
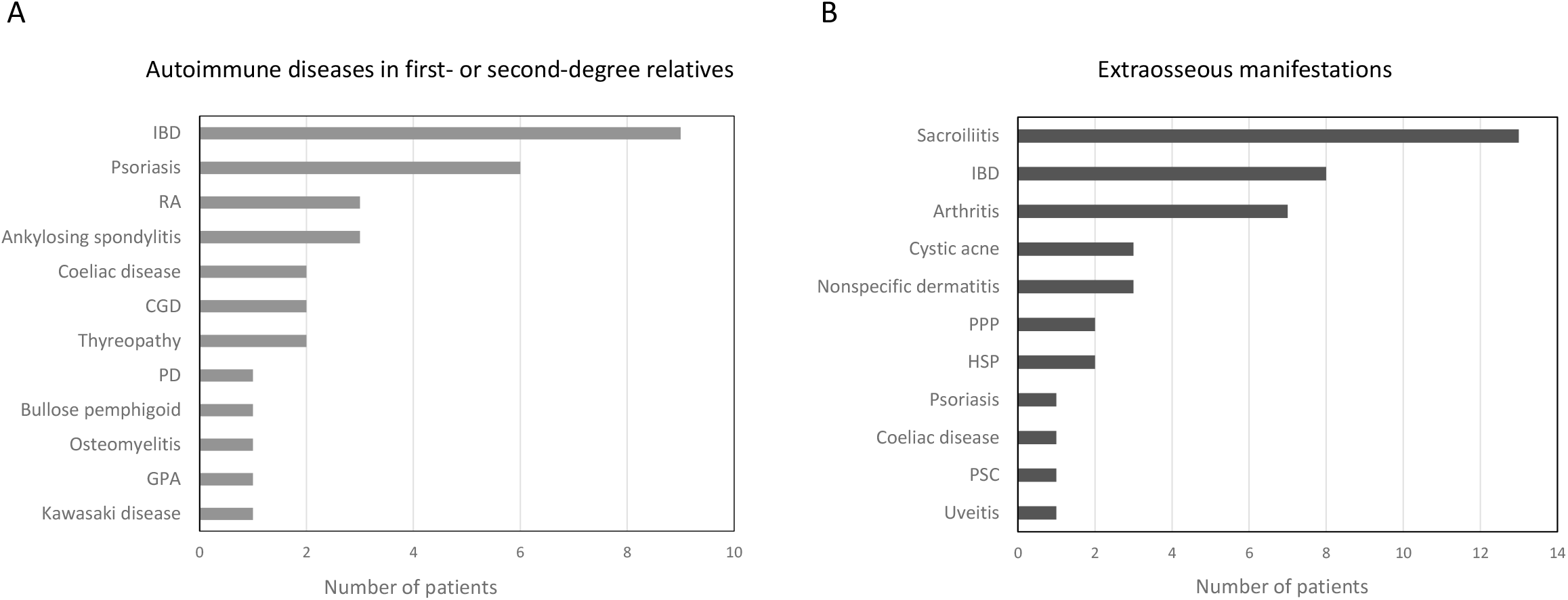
Autoimmunity profiles. **A)** Summary of total autoimmune disease manifestations in the first-and second-degree relatives and of **B)** extraosseous comorbidities in CRMO patients. CGD – chronic granulomatous disease, GPA – granulomatosis with polyangiitis, HSP – Henoch-Schönlein purpura, IBD – inflammatory bowel disease, PD – phagocytic dysfunction, PPP – palmoplantar pustulosis, PSC – primary biliary cirrhosis RA – rheumatoid arthritis

**Figure 3.**
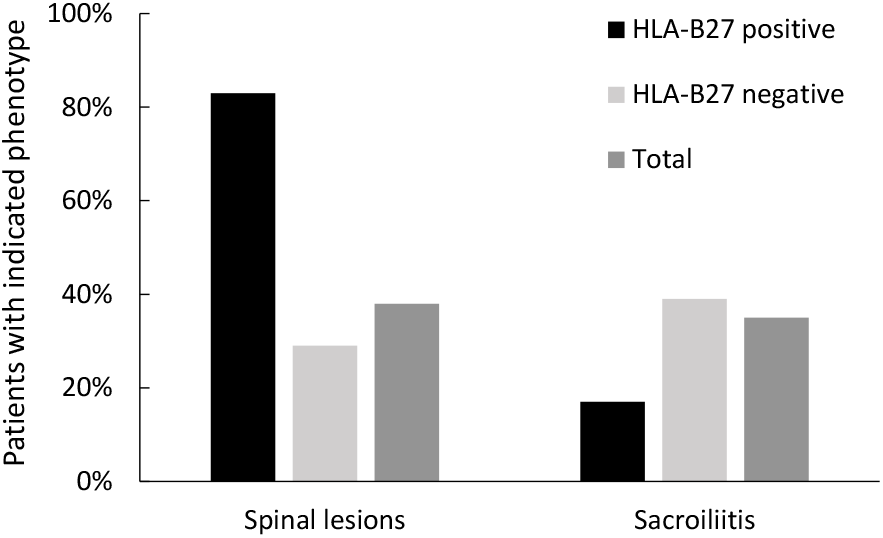
Vertebral and sacroiliac involvement. HLA-B27 positive children with CRMO displayed sacroiliac involvement less frequently than spinal involvement. Vertebral lesions were twice as frequent as in the cohort. Sacroiliitis in HLA-B27 positive CRMO was less frequent than sacroiliitis in HLA-B27 negative CRMO children and in non-selected CRMO children.

Eight children in our cohort (22%) developed enteropathic CRMO, osteomyelitis associated with IBD. Four had family histories of IBD. Tab. 2 summarizes characteristic features and the sequential course of their disease. CRMO preceded IBD in 3 patients and IBD preceded CRMO in 5. The average delay between presentation and manifestation of the associated disorder was 4 years. Two children who initially presented with CRMO and in whom complete remission of bone disease was achieved later developed IBD without relapse of CRMO. Enteropathic CRMO appears to be a specific subtype of the disease with a more severe phenotype. Such children in our study frequently suffered from more than one extraosseous manifestation, including skin disease, sacroiliitis, arthritis, primary sclerosing cholangitis, and uveitis. All children with enteropathic CRMO had elevated faecal calprotectin levels (Tab. 2). Mild elevation of faecal calprotectin (FCP; up to 250 μg/g) was found in samples from 9 other CRMO patients, but they did not progress to IBD, and their FCP levels dropped during follow-up.

Arthritis was diagnosed in 7 children (19%); 6 of them had a family history of autoimmune disease. Arthritis primarily but not exclusively adjoined foci of osteomyelitis. Eight patients (22%) developed skin disease including acne (n=3), pustular psoriasis (n=1), palmoplantar pustulosis (n=2), and atopic or unspecified dermatitis (n=3). SAPHO syndrome was diagnosed in 3 individuals (8%) based on severe nodulocystic acne and clavicular involvement with typical imaging findings. All 3 patients had an inflammatory phenotype at the time of diagnosis. Palmoplantar pustulosis occurred as a paradoxical effect of TNFi therapy (1x infliximab, 1x adalimumab) in 2 children who initially suffered from IBD and later manifested CRMO. Their skin disease was managed by topical treatment without withdrawal of TNFi therapy. One patient presented with chronic remitting pustular dermatitis but was not diagnosed with psoriasis. Her skin disease continues into adulthood. Two different patients with enteropathic CRMO had atopic dermatitis and nail pitting, but not skin disease corresponding to psoriasis.

In the next part of our study, we aimed to compare solely osteolytic and complex CRMO in children. A family history of autoimmune diseases was present in 71% of children with osteolytic CRMO and in 65% of children with complex CRMO. No differences were found for blood markers like leukocyte and platelet counts, haemoglobin, and mean cell volume (MCV) (Fig. 4A). By contrast, inflammatory activity was significantly higher in complex than in osteolytic disease (CRP, p=0.018; ESR, p=0.0064; Fig. 4B). Children with osteolytic CRMO manifested disease slightly earlier (mean 105 ± 36 months) than did children with complex CRMO (mean 123 ± 38 months); however, the difference was statistically not significant (p=0.1336). Nor did the number of lesions differ between the two groups (Fig. 4B). Interestingly, we found statistically significant differences in affected sites between the two groups. Whereas children with osteolytic CRMO often displayed long-bone involvement, children with complex CRMO presented predominantly with involvement of clavicle and pelvis (clavicle, p=0.011; pelvis, p=0.038; Fig. 5). Frequent and almost equal between both patient groups was femoral (50% in osteolytic CRMO, 48% in complex CRMO) and spinal involvement (43% in osteolytic CRMO, 35% in complex CRMO; see also Tab. 3).

**Figure 4.**
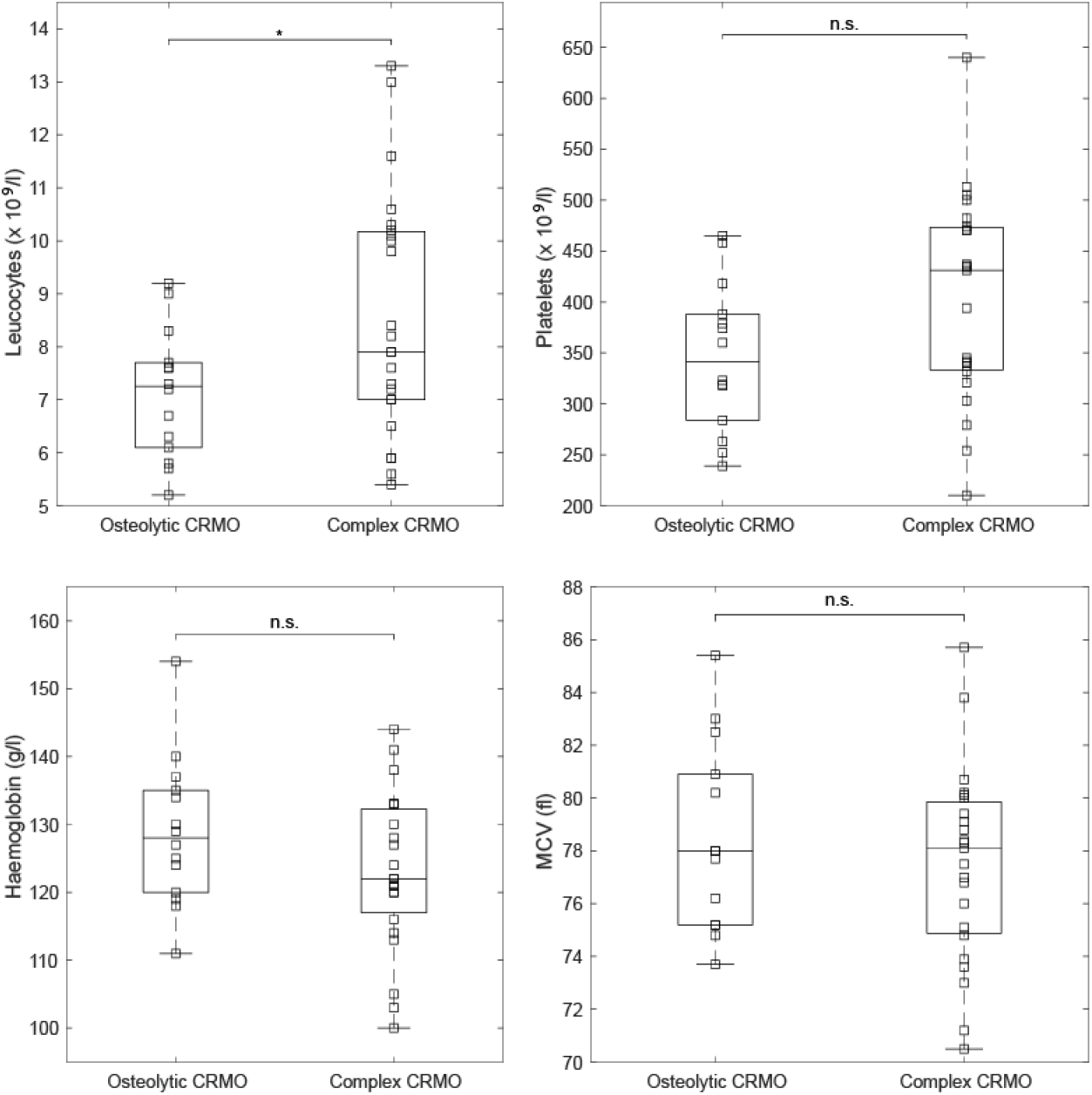
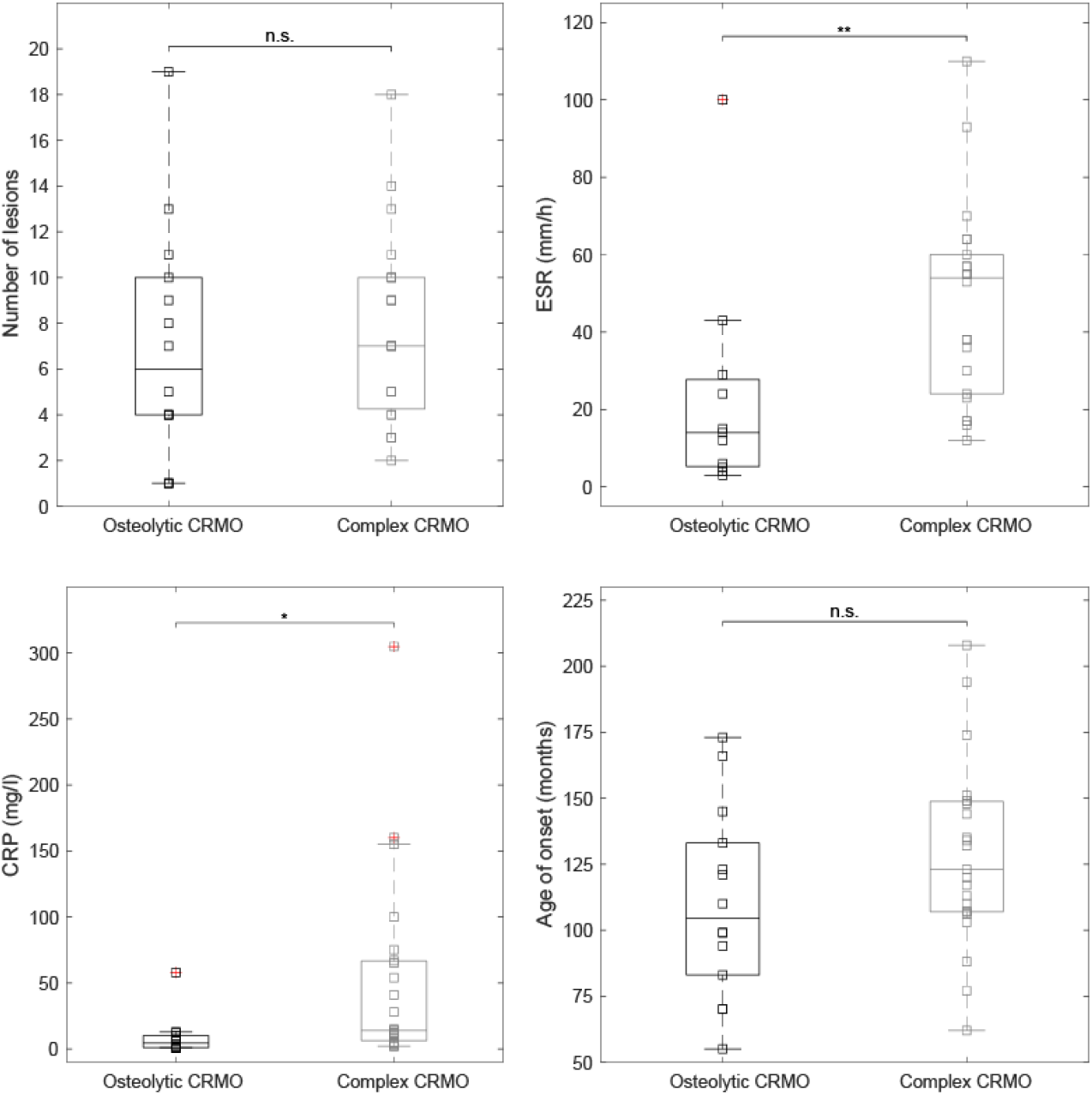
Comparison of laboratory characteristics between osteolytic and complex CRMO patients. A. Charts comparing blood markers, white blood count, platelets, haemoglobin, and mean cell volume (MCV) for the two groups of CRMO patients. Data are presented as boxplots with the 25th, 50th (median), and 75th percentiles marked. Whiskers extend to the most extreme data points. Squares represent individual patients. An asterisk (*) indicates statistical significance (p<0.05). **B**. Comparison of inflammatory markers, number of lesions, and age at onset between osteolytic and complex CRMO. Data are presented as boxplots with the 25th, 50th (median), and 75th percentiles marked. Whiskers extend to the most extreme data points. Squares represent individual patients. An asterisk (*) and two asterisks (**) indicate statistical significance (p<0.05 and p<0.01, respectively).

**Figure 5.**
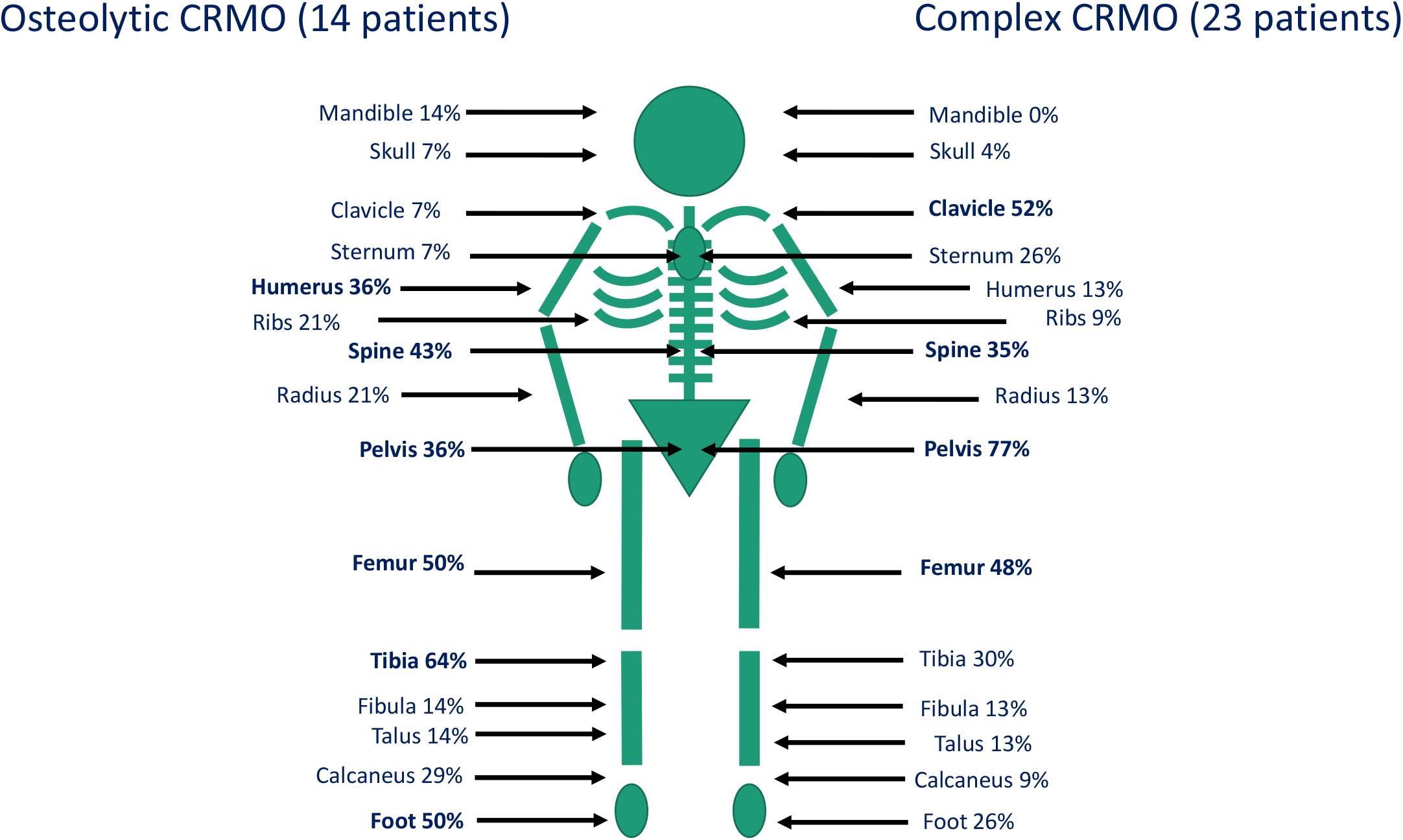
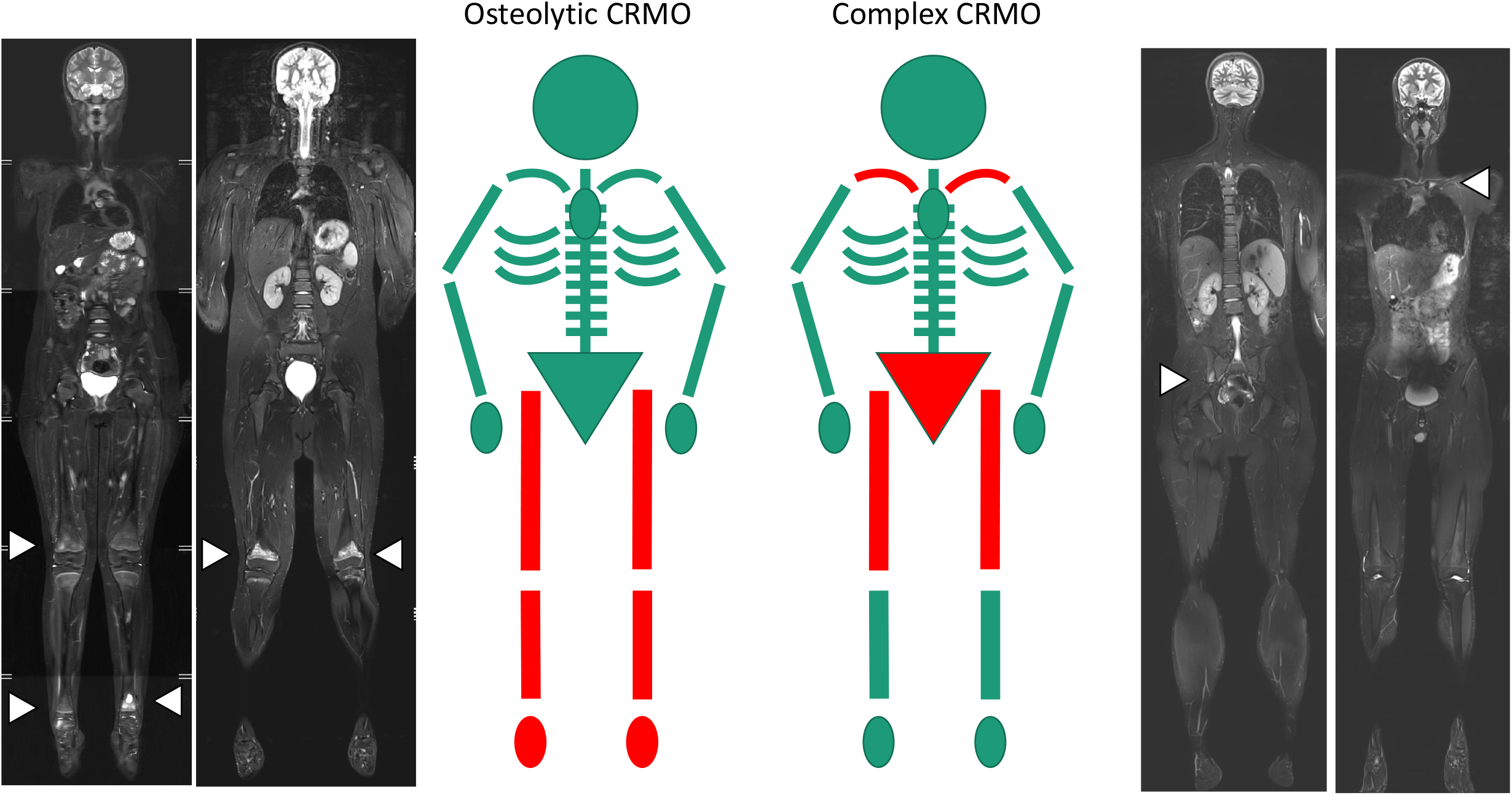
Distribution of CRMO lesions in osteolytic CMRO patients (left) and complex CRMO patients (right). A. Frequencies ≥ 35% are considered important (bold). P-values for each site are summarized in Table 3. **B**. Dominant patterns (red) of affected bones in osteolytic CMRO (left) and complex CRMO (right) with illustrative cases of WB-MRI. Arrowheads indicate main affected sites.

**Table 1.** Demographic and clinical characteristics of the cohort.

|  |  |
| --- | --- |
| **Total number of patients** | **37** |
| **Female** | 30 (81%) |
| **Male** | 7 (19%) |
| **Ethnicity** |  |
| - Romani | 4 (11%) |
| - Other Czech | 33 (89%) |
| **Age at onset, years; median (range)** | 10 (4.6 – 17.3) |
| **Number of lesions; median (range)** | 7 (1-19) |
| **Skeletal involvement** |  |
| - Unifocal | 3 (8%) |
| - Multifocal | 34 (92%) |
| **Initial symptom** |  |
| - Back pain | 14 (38%) |
| - Limb pain | 19 (51%) |
| - Other | 4 (11%) |
| **Bone biopsy** | 23 (62%) |
| **Spinal lesions** | 14 (38%) |
| - Spinal surgery | 7 (19%) |
| **Sacroiliac joint involvement** | 13 (35%) |
| **Inflammatory bowel disease** | 8 (22%) |
| **Psoriasis / palmoplantar pustulosis** | 3 (8%) |
| **Nodulocystic acne** | 3 (8%) |
| **Unspecific dermatitis involvement** | 3 (8%) |
| **Autoimmune comorbidities in family history** | 25 (68%) |
| - First-degree (I) relatives | 15 |
| - Second-degree (II) relatives | 13 |
| **Anti-nuclear antibodies present** | 2 (5%) |
| **HLA-B27 present** | 6 (16%) |

**Table 2.** Characteristics of patients with enteropathic chronic recurrent multifocal osteomyelitis.

| No. | Gender | No. foci | Autoantibodies | Histology | Disease course | Delay (years) | Therapy | Outcome | HLA-B27 | FCP (µg/g) | Family history |
| --- | --- | --- | --- | --- | --- | --- | --- | --- | --- | --- | --- |
| 1. | F | 7 | ASCA positive | NS colitis | 1. CRMO, arthritis<br>2. colitis + CRMO relapse | 5.0 | SSZ --> P-->ADA+MTX --> none | remission, now without therapy | negative | 1000 | II. HLA-B27 AS |
| 2. | M | 10 | p-ANCA positive | CD | 1. IBD<br>2. PPP<br>3. CRMO + IBD relapse | 4.0 | AZA, IFX, IFX+MTX -->ADA | relapsing IBD and CRMO | negative | 1736 | I. IBD |
| 3. | F | 2 | ASCA positive | CD | 1. CRMO, arthritis<br>2. IBD<br>3. HSP<br>4. acute uveitis | 0.5 | SSZ --> ETA+MTX --> ADA, AZA | remission on therapy | positive | 350 | I. IBD |
| 4. | F | 9 | ASCA positive | CD | 1. IBD<br>2. CRMO+PPP | 4.0 | AZA -->IFX-->ADA-->ADA+SSZ-->ADA+MTX+SSZ | remission on therapy | negative | 3329 | II. psoriasis |
| 5. | F | 3 | p-ANCA positive | UC | 1. IBD<br>2. relapsed IBD+CRMO | 2.5 | P+AZA-->5-ASA-->ADA+MTX | remission on therapy | negative | 938 | negative |
| 6. | F | 11 | negative | UC, PSC | 1. IBD, PSC<br>2. CRMO | 4.0 | 5-ASA, UDCA, PAM-->ADA | active CRMO, IBD in remission | negative | 1680 | II. IBD |
| 7. | M | 7 | p-ANCA positive | CD | 1. CRMO, arthritis<br>2. IBD (arthritis and CRMO in remission) | 7.5 | NSAID--> SSZ, P, MTX --> ADA | active IBD, CRMO in remission | negative | 1230 | unknown |
| 8. | M | 9 | negative | CD | 1. CRMO<br>2. IBD (CRMO in remission) | 4.0 | NSAID--> P-->5-ASA-->non-compliance | active IBD, CRMO in remission | negative | 545 | I. IBD, II. psoriasis |

Abbreviations: ADA – adalimumab, ANCA – anti-neutrophil cytoplasmic antibody, ASCA – anti-*Saccharomyces cerevisiae* antibody, 5-ASA – 5-aminosalicylic acid, AZA – azathioprine, CD – Crohn disease, CRMO – chronic recurrent multifocal osteomyelitis, ETA – etanercept, FCP – foetal calprotectin, I. – first-degree relative, II. – second-degree relative, HLA-B27 – human leucocyte antigen B27, HLA-B27 AS – HLA-B27 positive ankylosing spondylitis, IBD – inflammatory bowel disease, IFX – infliximab, MTX – methotrexate, NS colitis – non-specific colitis, NSAID – non-steroidal anti-inflammatory drug, P – prednisone, PAM – pamidronate, PPP – palmo-plantar pustulosis, PSC – primary sclerosing cholangitis, SSZ – sulphasalazine, UC – ulcerative colitis, UDCA – ursodeoxycholic acid.

**Table 3.** Distribution of lesions in patients with osteolytic and complex CRMO.

| Affected site | Osteolytic CRMO<br>(no = 14) | Complex CRMO<br>(n = 23) | P value |
| --- | --- | --- | --- |
| skull | 1 | 1 | 1.000 |
| mandible | 2 | 0 | 0.137 |
| scapula | 1 | 1 | 1.000 |
| clavicle | 1 | 12 | 0.011 |
| sternum | 1 | 6 | 0.217 |
| humerus | 5 | 3 | 0.215 |
| radius | 3 | 3 | 0.653 |
| ribs | 3 | 2 | 0.346 |
| spine | 6 | 8 | 0.732 |
| pelvis (incl. sacrum) | 5 | 17 | 0.038 |
| femur | 7 | 11 | 1.000 |
| tibia | 9 | 7 | 0.086 |
| fibula | 2 | 3 | 1.000 |
| talus | 2 | 3 | 1.000 |
| calcaneus | 4 | 2 | 0.173 |
| feet with digits | 7 | 6 | 0.171 |

Treatment options used in our cohort are summarized in Fig. 6. All children with osteolytic CRMO and 74% of those with complex CRMO received NSAIDs, including ibuprofen, naproxen, or diclofenac. Half of the patients were treated with bisphosphonates (pamidronate or zoledronate) because of spinal involvement or inefficacy of NSAIDs. The percentage of bisphosphonate treatment was similar for the osteolytic and complex CRMO groups (respectively 43% and 50%), while spinal involvement was identified in respectively 43% and 35% of children. Children with complex CRMO more often received conventional synthetic DMARDs, corticosteroids, and biologic therapy (TNFi) (Fig. 6). Substantial improvement or disease remission was achieved in 27 patients (73%); 7 children (19%) still have active chronic or remittent disease. Three patients with complex CRMO were lost to follow-up due to non-compliance.

**Figure 6.**
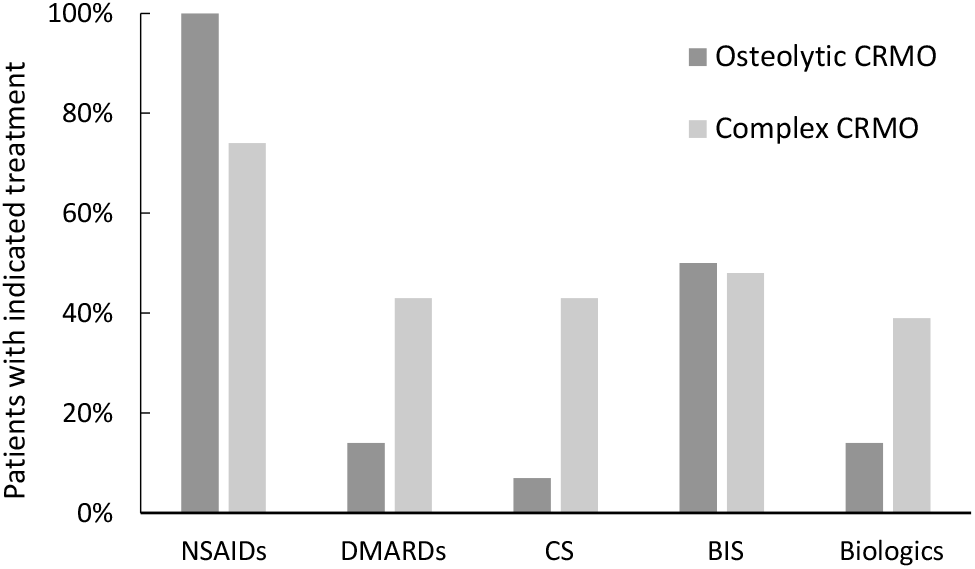
Treatments applied in patients with osteolytic (dark grey) and complex (light grey) CRMO. NSAID - non-steroidal anti-inflammatory drugs, DMARDs – disease modifying antirheumatic drugs, CS – corticosteroids, BIS – bisphosphonates, TNFi – tumour necrosis factor α inhibitors

## Discussion

In this study, we focused on different phenotypes of CRMO. We observed that children with extraosseous involvement present with significantly increased inflammatory activity, have a distinct type of bone involvement, and more frequently require escalation to a second-line therapy. No differences in number of bone lesions and in age at disease onset were found.

Some earlier studies noticed phenotypic differences between CRMO patients and endeavoured to stratify their patients according to severity and disease course ^7,17,25^. Wippf et al. defined three subgroups of CRMO patients ^7^. Compared to the mild form, children with intermediate and severe phenotypes more frequently presented with extraskeletal co-morbidities, multifocal disease, and inflammatory syndrome. The more aggressive CRMO phenotype, with higher rates of multifocality, higher ESR, and predominant involvement of feet, tibia, pelvis, and humerus was described in children with extraosseous manifestation by Borzutzky and colleagues ^6^. In accordance with these findings, O’Leary et al. demonstrated multifocality of lesions and low response rate to NSAIDs in their Irish cohort with a high prevalence of extraosseous involvement. We observed similar differences between children with and without extraosseous manifestations. Our findings further suggest that lesions in complex CRMO patients with affected extraskeletal tissues, particularly skin, gut, and joints, are differently distributed. Clavicular and pelvic lesions dominated in complex CRMO whereas lesions in metaphysis of long bones and foot were less frequent (Fig. 5B). On the contrary, foot and long bones, specifically tibia, were prevalent sites of lesions in osteolytic CRMO. In our group of patients, we noticed an unusually high incidence of IBD compared with other cohorts. In the subgroup of patients with IBD, the dominant osteolytic pattern comprises clavicle and pelvis. However, further studies on a large cohort are required for generalisation of our observations and determination of predictive value of lesion distribution in CRMO subtypes.

Family history has an important, yet not fully understood role in CRMO. More than 50% of children in reported cohorts had family histories of autoimmune disorders, especially of those on the spondyloarthritis spectrum. One of the characteristics of these disorders, recently extensively studied, is the alteration of gut homeostasis and of the enterobiome. Extraosseous manifestations observed in our patients and autoimmune diseases in families exhibit similarities (Fig. 2). Arthritis, IBD, and psoriasis are conditions shared between the two groups. However, specific autoimmune diseases are not clustered within the families in any apparent pattern. For example, only half of the children with enteropathic CRMO in our cohort have a family history of IBD. On the other hand, arthritis or psoriasis, also present in pedigrees of our patients, is known to be associated with alteration of gut microbiota ^26-29^. Thus, it can represent an additional factor contributing to the development of enteropathic CRMO. Indeed, such correlation was proposed in two CRMO studies ^27, 30^. A recent report also describes successful deployment of a Crohn-disease exclusion diet in a paediatric patient with enteropathic CRMO ^31^. Interestingly, we observed elevated FCP levels in 31% of our patients with active CRMO (apart from those with the enteropathic form), indicating the presence of subclinical gut inflammation. The retrospective character of this study prevented analysis of the microbiome in our patients. Nevertheless, a better understanding of the interplay between genetic predispositions, mirrored in family history of the probands, and environmental factors could promote our understanding of disease aetiopathogenesis and possibly shape future therapy strategies. A possible inference is that children with a higher risk of IBD may profit from an earlier switch to TNFi or from NSAIDs avoidance. Similarly, to identify patients at high risk for development of psoriasis, including paradoxical disease during TNFi treatment, could encourage concomitant treatment with DMARDS, such as methotrexate ^32,33^.

Even though our study has yielded several interesting findings, we must mention limitations that restrain us from offering general recommendations. First and foremost, the number of children with CRMO in this cohort is low. The retrospective approach in this study did not allow implementation of a uniform treatment protocol for all patients and a comprehensive evaluation of the treatment response. Importantly, our findings are in line with other studies indicating that CRMO is a heterogeneous disease ^34,35^ and that “CMRO” might be considered as an umbrella term for phenotypically similar disorders with potentially diverse etiopathogenetic backgrounds and sterile osteomyelitis as a dominant feature.

In conclusion, osteomyelitis as a dominant phenotypic feature of CRMO is accompanied by inflammation of other tissues in a substantial group of patients. The disease thus presents in two main forms, osteolytic and complex. The concept of CRMO as a clinical continuum between these two distinctive forms impedes definition of diagnostic and therapeutic strategies. A tailored therapy of CRMO patients remains unavailable. However, a detailed clinical stratification of patients followed by genetic-epigenetic studies can promote early identification of high-risk patients, who could benefit from more aggressive treatment with early therapy escalation.

## Data Availability

All data produced in the present study are available upon reasonable request to the authors.

## Availability of data and materials

The datasets used and/or analysed during the current study are available from the corresponding author on reasonable request.

## Abbreviations

CNO: chronic non-bacterial osteomyelitis
CRMO: chronic recurrent multifocal osteomyelitis
CRP: C-reactive protein
DIRA: deficiency of interleukin-1 receptor antagonist
DMARD: disease-modifying anti-rheumatic drug
FCP: foetal calprotectin
HLA: human leukocyte antigen
IBD: inflammatory bowel disease
MRI: magnetic resonance imaging
NSAID: non-steroidal anti-inflammatory drug
SAPHO: synovitis, acne, pustulosis, hyperostosis, osteitis
TNFi: tumour necrosis factor inhibitor
WB-MRI: whole-body MRI

## Acknowledgement

We would like to thank A. S. Knisely (Med Univ Graz, Austria) for critical comments and revision of the manuscript.

## Author information Contributions

DC designed the study, provided patient information and wrote the manuscript, HM provided patient information and reviewed manuscript, VK re-assessed X-ray and WBMR imaging and reviewed manuscript, ZK performed statistical analyses, OS contributed to data analyses and reviewed the manuscript, LW provided patient data and reviewed the manuscript, JB contributed to data analyses and reviewed the manuscript, ZŠ contributed to data analyses and reviewed the manuscript, MK re-assessed X-ray and WBMR imaging, provided WBMR pictures for Fig. 5B and reviewed manuscript, MC contributed to the critical revisions of manuscript, data analyses and prepared the figures and tables, RH contributed to data analyses and reviewed the manuscript. All authors agreed with final version of the manuscript.

## Corresponding author

Correspondence to Dita Cebecauerova.

## Ethics declarations

### Ethics approval and consent to participate

All participants or their legal guardians gave written consent in participating in the study. The study was performed as an retrospective cohort study according to the principles of the Declaration of Helsinki and was approved by the local ethics committee at the University Hospital Motol (protocol No. EK 1412/20).

